# COVID-19 Preprints and Their Publishing Rate: An Improved Method

**DOI:** 10.1101/2020.09.04.20188771

**Authors:** Francois Lachapelle

## Abstract

**Context:** As the COVID-19 pandemic persists around the world, the scientific community continues to produce and circulate knowledge on the deadly disease at an unprecedented rate. During the early stage of the pandemic, preprints represented nearly 40% of all English-language COVID-19 scientific corpus (6, 000+ preprints | 16, 000+ articles). As of mid-August 2020, that proportion dropped to around 28% (13, 000+ preprints | 49, 000+ articles). Nevertheless, preprint servers remain a key engine in the efficient dissemination of scientific work on this infectious disease. But, giving the ‘uncertified’ nature of the scientific manuscripts curated on preprint repositories, their integration to the global ecosystem of scientific communication is not without creating serious tensions. This is especially the case for biomedical knowledge since the dissemination of bad science can have widespread societal consequences.

**Objective:** In this paper, I propose a robust method that allows the repeated monitoring and measuring of COVID-19 preprints’ publication rate. I also introduce a new API called *Upload-or-Publish*. It is a free micro-API service that enables a client to query a specific preprint manuscript’s publication status and associated meta-data using a unique ID. The beta-version is currently working and deployed.

**Data:** I use Covid-19 Open Research Dataset (CORD-19) to calculate COVID-19 preprint corpus’ conversion rate to peer-reviewed articles. CORD-19 dataset includes 10,454 preprints from arXiv, bioRxiv, and medRxiv.

**Methods:** I utilize conditional fuzzy logic to link preprints with their published counterparts. My approach is an important departure from previous studies that rely exclusively on bio/medRxiv API to ascertain preprints’ publication status.

**Results:** As expected, the findings suggest a positive relationship between the time elapsed since preprints’ first server upload and preprints harboring a published status. For instance, as of mid-September, close to 50% of preprints uploaded in January were published in peer-review venues. That figure is at 29% for preprints uploaded in April, and 5% for preprints uploaded in August. As this is an ongoing project, it will continue to track the publication rates of preprints over time.

## Introduction

In their article Preprinting a Pandemic, Fraser et al. (2020) document the unparalleled role of preprints and the digital repositories hosting those scientific manuscripts in the diffusion of COVID-19 science. Between January and April 2020, preprints accounted for nearly 40% of all English-language COVID-19 scientific work (Fraser et al., 2020). The current estimate puts that figure at 28% (NIH iSearch, 2020). Majumder and Mandl (2020:30) suggested that during the early days of the pandemic “because of the speed of their release, preprints – rather than peer-reviewed literature in the same topic area—might [have been what drove the] discourse related to the ongoing COVID-19 outbreak”. Speed and efficiency in the race to share and disseminate scientific knowledge about the disease is a clear reason for the massive adoption of preprint servers during the pandemic (Chiarelli, Johnson, Pinfield, and Richens, 2020). For instance, once submitted on its server, bioRxiv usually only takes two days to screen and upload a manuscript while medRxiv takes four days (Fraser et al., 2020; Abdill & Blekhman, 2019a; Kwon, 2020). In addition to transforming how scientists share their work, preprint servers in the context of the pandemic are also changing the form-factor of knowledge itself as preprint manuscripts tend to be significantly shorter, use fewer references and tables (Fraser et al., 2020; Lin et al., 2020). But, giving the ‘uncertified’ nature of the scientific manuscripts curated on preprint repositories, their integration to the global ecosystem of scientific communication is not without creating serious tensions, a double-edged sword of sort. This is especially the case for biomedical knowledge since the dissemination of bad science can have widespread consequences for a host of health practitioners and social actors who changed their professional practices or/and behaviors based on scientific findings. The case of chloroquine and hydroxychloroquine as a viable treatment to COVID-19 is a clear example of the risks of circulating bad or unascertained science (Pastick et al., 2020).

And indeed, in the past few months, several initiatives have emerged to mitigate this inherent problem of preprint research’s unreviewed-ness. In early March, Nature announced the launch of Outbreak Science Rapid PREreview, a platform “for rapid review of preprints related to emerging outbreaks” (OutbreakScience, 2020). Even more recently MIT introduces Rapid Reviews COVID-19 (RR:C19), a tool relying on AI and humans to identify important preprint manuscripts in need of peer-review (RR:C19, 2020). On the integration front, biomedical bibliometric powerhouses began indexing preprint manuscripts’ metadata to their platforms. For example, the National Library of Medicine (NLM) just launched the *NIH preprint pilot* with the goal indexing NIH-funded preprints on both PubMed Central (PMC) and PubMed websites (NIH preprint pilot, 2020). More than two decades ago, NIH’s first attempt to index preprints on PMC (Varmus, 1999; Smaglik, 1999) was scrapped when the National Academy of Sciences “successfully negotiated the exclusion of work that had not been peer-reviewed” (Abdill et al., 2019a: 3). Just as CORD-19 data science project has been doing since its inception in March 2020 (Wang et al., 2020), NIH also begin including preprint manuscripts from six sources in its curated COVID-19 online publication database, NIH iSearch COVID-19 portfolio (NIH iSearch, 2020).

Existing peer-reviewed journals are also participating in that integration push. Let take the example of eLife Celebrating preprint servers as “one of the most inspiring things about research in the age of COVID-19”, the journal for the biomedical and life sciences announced they made “posting to bioRxiv or medRxiv – either by the authors themselves or the journal – the default for all eLife submissions” (Eisen, Akhmanova, Behrens, and Weigel, 2020). Recognizing the utmost importance for the swift dissemination of knowledge in a time of global health-crisis, several medical journals have conducted internal reforms to their publication process. As a result, research indicates that the time between the submission and the publication of a manuscript has shrunk on average by 49% for coronavirus-related research (Horbach, 2020). The open-access nature of online repositories also highlighted the proprietary nature of academic publishing. By mid-March, after an online campaign by the academic community, more than 30 scientific publishers have decided to make all COVID-19 work freely accessible (Nature, 2020; Karr, 2020). Here, there is little doubt that preprint servers played a role in that decision to make knowledge more available. The journal PLOS has an option that can automatically upload a manuscript submitted on its website to bioRxiv, if the authors wish so.

Meanwhile, in an environment where publication norms are changing quickly and call for reforms in the peer-review system are mounting (Shopovski and Slobada, 2020), preprint repositories are also revising and tightening some of their screening policies. For example, early in the pandemic, bioRxiv announced that it “would no longer accept manuscripts making predictions about treatments for COVID-19 solely on the basis of computational work” (Kwon, 2020). In some corners of academia, we have preprint advocates defending preprint servers as a parallel system, that is, a potential and viable alternative model of knowledge communication to the traditional peer-review pipeline. Put differently, in fields like mathematics or biology, scholars have, in the past, opted to use preprint servers as the only and final venue to ‘publish’ their important work (Lariviere et al., 2014; Singh, 2017; Nature Bio, 2020). Studies—both on non-COVID-19 and COVID-19-related—have found evidence against the idea that preprint research doesn’t meet the gold-standard of peer-review research (Vale, 2015; Klein et al., 2016; Nature Bio, 2017; Fraser et al., 2020).

In the life science, however, “the potential harm from posting erroneous provisional research is one reason why the medical community was so cautious about preprints in the first place” (Gianola, Jesus, Bargeri, and Castellini, 2020:17; see also, Kwon, 2020; Cobb, 2017; Desjardins-Proulx et al. 2013; Abdill and Blekhman, 2019). The formal peer-review process that stands as the cornerstone of modern science can still intervene and judge preprint manuscripts after they are uploaded on digital repositories. bioRxiv server, for instance, is already seemingly aligned with the traditional peer-review system as more than 100 journals accept submissions directly from their repository (bioRxiv submission guide, 2020). Once accepted on its server, bioRxiv also automatically sends an email to the corresponding author to offer its help in getting the manuscript into the peer-review grinder. In other words, the major biomedical preprint server was already quite integrated to the formal scholarly channel before the beginning of the pandemic. Preprint servers might seem to operate as a parallel system in the midst of the pandemic, but the landing page of the major biomedical repositories’ website continue to stipulate that preprints are not peer-reviewed articles – “[Preprints] should not be relied on to guide clinical practice or health-related behavior” (medRxiv landing page, 2020).

Findings on publishing practices during the early phase of the pandemic indicate that only 8.6% (n=329) of the 3,805 medRxiv and bioRxiv COVID-19 preprints uploaded up to May 20, 2020 were published in peer-reviewed venues (Gianola et al., 2020). The only other research measuring medRxiv and bioRxiv COVID-19 preprints uploaded up to April 31^st^, 2020 report a publishing rate of 4% (n=101) (Fraser et al., 2020). As for studies on the fate of preprints on the peer-review pipeline unrelated to COVID-19, upon reviewing the emerging literature, Abdill and Bleckhman (2019a: 26-27) conclude that “the majority of bioRxiv preprints do appear in journals”. For instance, if Schloss (2017) found that only 33.6 % of bioRxiv manuscripts uploaded before 2015 were published in peer-review journals, Abdill et al. (2019a) who reproduce a similar study in 2018 report that close to 70% have been published. More recently, Lin, Yu, Zhou, Zhou, and Shi (2020) found that nearly 80% of computer science (CS) related preprints uploaded to the arXiv repository server between 2008 and 2017 were eventually published in peer-reviewed venues. Time is an important dimension as a manuscript need time to make their way through the peer-review system. For bioRxiv, findings indicate 75% of published preprints appears in a journal within eight months of their initial upload (Abdill et al., 2019a:27). For arXiv, this figure is about 12 months (Lariviere et al., 2014). This is also what the only two COVID-19 studies of preprints’ conversion rate suggest. Between April and May 2020, the proportion of published preprints appears to have double.

### Objectives

Given the strong integration movement described earlier and research indicating the positive relationship between time elapse since first upload and formal publication it is likely that a reproduction of Gianola et al.’s or Fraser et al.’ study will find an increased proportion of published COVID-19 preprints. That said, the core objective of this paper is to introduce a more robust method to measure COVID-19 preprints’ conversion rate. Both studies just discussed rely on bioRxiv/medRxiv API to ascertain the publishing status of COVID-19 preprints. But recent findings indicate that the proportion of false negatives unpublished preprints that were in fact published – to be as high as 37.5% (Abdill et al., 2019a:15). Therefore, preprint repositories API metadata should be used only as a supplementary method and not as the primary means to determine preprints’ publication status. The objectives of this paper are three-fold:

1. First, to provide an updated measure of COVID-19 preprints’ ‘conversion rate’. In this paper, using CORD-19 data and conditional fuzzy matching, I measure the proportion of preprints produce between January and September 2020 have been published as peer-reviewed articles, a measure I will refer to as ‘conversion rate’.
2. This paper is more than a ‘replication study’. I propose a robust steam-data system that can automatically calculate the COVID-19 preprints’ conversion rate every time CORD-19 updated their dataset which is usually twice a week. In other words, I develop a Python program that ensures not only the reproducibility but also the frequent monitoring of this important metric.
3. Last, but not least, I introduce a new API called *Upload-or-Publish*. Also written using Python programming language, the micro-API service allows a client to query a specific preprint manuscript using a unique ID—DOI parameter for beta-version. The API returns a JSON dictionary first indicating if the preprint in question has been published as a peer-reviewed article. If so, the API returns additional metadata of the published articles (pr_title, pr_journal, pr_date, pr_author). The data is also available in a database format. I am confident this tool will join an exciting pool of data science tools—Rxivist.org (Abdill et al., 2019b), bioPreprint (Iwema, LaDue, Zack, and Chattopadhyay 2016)—that aims at (1) providing accurate and new data-points pertaining to the preprint environment and (2) laying the groundwork for more in-depth empirical examinations of preprint practices.

### CORD-19 Dataset

I use Covid-19 Open Research Dataset (CORD-19) to calculate COVID-19 preprint corpus’ conversion rate to peer-reviewed articles. Arguably the most ambitious bibliometric COVID-19 project, CORD-19 is the collaborative effort between the Allen Institute for AI and half a dozen organizations including NIH and the White House (for more details, see Wang et al., 2020). CORD-19 was built with data scientists in mind as its main user base as it was “designed to facilitate the development of text mining and information retrieval systems” (Wang et al. 2020:1). What makes CORD-19 stand-out in the always-growing field of COVID-19 bibliometric projects is the fact that in addition to providing the largest dataset containing over 250, 000 coronaviruses-related publications’ metadata, CORD-19’s freely downloadable zip file includes over 87,000 fully parsed full text.

### Early Inclusion of Preprints

CORD-19 is a record-linkage project that merges coronaviruses-related publications from Elsevier, PMC, WHO, PubLine, and three preprint servers—arXiv, bioRxiv, and medRxiv. To the author’s knowledge, before NIH launched its own COVID-19 bibliometric platform (NIH iSearch), CORD-19 was the only project to include English-language COVID-19-related preprint manuscripts to its database. The curation of preprints’ metadata (author(s), title, venue, etc.) in the dataset alongside peer-reviewed publications’ is what allows us to use a string matching technique –fuzzy logic– to identify which preprints have been published in peer-reviewed venues.

NIH iSearch includes a variable called “Published As” which indicates COVID-19 preprints’ publication status. But my extensive data exploration reveals that NIH iSearch’s variable is not reliable enough to replace a systematic record-linkage effort on COVID-19 population of publications. And for two reasons. First, NIH relies on bio/medRxiv API to determine preprints’ publication status. This is problematic since bio/medRxiv API’s level of false negatives—or preprints marked as unpublished that are published—could be as high as 37% (Abdill et al., 2019a). Secondly, NIH iSearch erroneously marked as “published” a large number of preprints that were simply uploaded on multiple online preprint repositories. In short, both type I and type II errors prevent us to use NIH iSearch to generate a reliable estimate of publication rate for the leading biomedical preprint servers.

### Filtering non-COVID-19 Publications

I conducted my analysis using the 49th version of the CORD-19 metadata dataset (retrieved on September 12^th^, 2020). The raw csv file (metadata.csv) contains 253, 454 publications. As already mentioned, since CORD-19 covers not only COVID-19 publications, but all coronaviruses-related literature published in English-language since the 1950s, I use two filtering strategies to identify COVID-19 articles. First, I exclude all articles with a publishing date prior 2019 since we know the first COVID-19 article was published that year. This step removed 33% of all cases, or 82,992 publications (see Table 1). In a second time, since not all 170,462 remaining cases with the publishing year of 2019 or 2020 are COVID-19 research, I utilize a keyword approach to identify COVID-19 related publications (for a systematic list, see Leaman, 2020). I relied on a curated list of 19 keywords to carry this second filtering step (see Table 2). For each case, if the abstract or title (CORD-19 dataset’s field or variable) contains at least one of the COVID-19 keywords, the Python program marks the case as a valid COVID-19 publication. This algorithmic step brought the dataset size to 122,152 as it retained 72% of the remaining cases. Using the keyword method on the NIH iSearch dataset (n=49116) as a validation set accurately identified 98% of the cases.

**Table 1.** Data Cleaning: Identification of COVID-19 Corpus in CORD-19

| **Cleaning Step** | **Description** | **Case Left**<br>**(n)** | **Excluded**<br>**(n)** | **Excluded**<br>**(%)** |
| --- | --- | --- | --- | --- |
| raw CORD dataset |  | 253454 | NA | NA |
| Filter 1 - year | excludes documents published before 2019 | 170462 | 82992 | 0.33 |
| Filter 2 - keyword | excludes non-COVID-19 documents | 122152 | 48310 | 0.19 |
| Filter 3 - language | excludes non-english lang. documents | 117464 | 4688 | 0.02 |
| Filter 4 - duplicate | excludes duplicate cases - published | 84852 | 32612 | 0.13 |
| Filter 5 - duplicate | excludes duplicate cases - preprint | 84607 | 245 | 0.00 |
| **Total** |  | **84607** | **168847** | **0.67** |

**Table 2.**
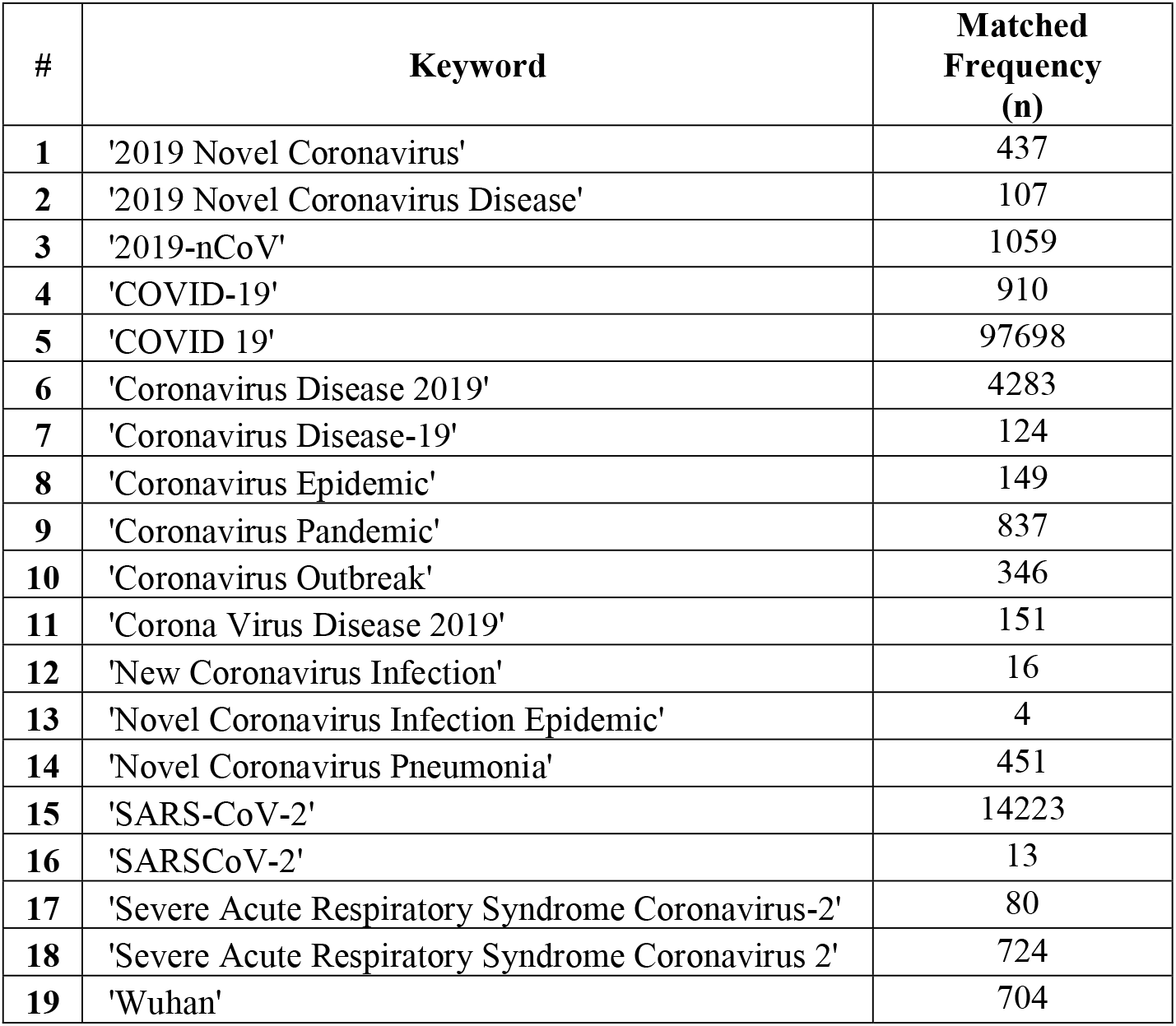
List of COVID-19/SARS-Cov-2 Keywords

| # | Keyword | Matched Frequency (n) |
| --- | --- | --- |
| 1 | '2019 Novel Coronavirus' | 437 |
| 2 | '2019 Novel Coronavirus Disease' | 107 |
| 3 | '2019-nCoV' | 1059 |
| 4 | 'COVID-19' | 910 |
| 5 | 'COVID 19' | 97698 |
| 6 | 'Coronavirus Disease 2019' | 4283 |
| 7 | 'Coronavirus Disease-19' | 124 |
| 8 | 'Coronavirus Epidemic' | 149 |
| 9 | 'Coronavirus Pandemic' | 837 |
| 10 | 'Coronavirus Outbreak' | 346 |
| 11 | 'Corona Virus Disease 2019' | 151 |
| 12 | 'New Coronavirus Infection' | 16 |
| 13 | 'Novel Coronavirus Infection Epidemic' | 4 |
| 14 | 'Novel Coronavirus Pneumonia' | 451 |
| 15 | 'SARS-CoV-2' | 14223 |
| 16 | 'SARSCoV-2' | 13 |
| 17 | 'Severe Acute Respiratory Syndrome Coronavirus-2' | 80 |
| 18 | 'Severe Acute Respiratory Syndrome Coronavirus 2' | 724 |
| 19 | 'Wuhan' | 704 |

### Duplicates

Because CORD-19 dataset includes both preprints and published work, filtering out duplicates requires special attention. It is not uncommon for a preprint and its peer-reviewed published counterpart to share the exact same title (Lin et al., 2020). Therefore, if we were to use ‘article title’ to identify case duplication in the CORD-19 publication dataset, we would effectively flag either several preprints or its published counterparts and remove them from the final dataset. In other words, it would prevent us to accurately measure the publication rate of preprints. Therefore, to avoid this problem, I first identified and removed duplicates amongst preprint titles before applying the same procedure for the published titles, or peer-reviewed documents. I found 245 duplicates among preprint documents and 32,612 duplicates (13%) among peer-reviewed articles. The final, clean CORD-19 dataset contains a total of 84,607 cases, or 33% of the original total number of cases.

### Preprints Documents in CORD-19 Dataset

In the final dataset I use for analysis, COVID-19 preprint manuscripts represent a little more than 12% of all publications indexed in CORD-19 dataset (n=10,454; 12.4%). That said, it is important to note that CORD-19 includes not only preprints and peer-reviewed articles but also other forms of scholarly communications such as book or volume chapters. Those scholarly documents can be tricky to identify and remove from the CORD-19 dataset. Therefore, to obtain a more accurate proportion of preprints to peer-reviewed documents, if one use LitCovid dataset’s total figure which only included English-language peer-reviewed documents, we found that arXiv and bio/medRxiv alone represents around 17% of all scientific work. According to Fraser et al. (2020), between January and April 2020, preprints accounted for nearly 40% of all English-language COVID-19 scientific work.

In terms of the source distribution, nearly two-thirds of all preprint articles found in CORD-19 dataset emanated from medRxiv (n=6,450; 62.5%), with bioRxiv (n=1,839; 17.5%) and arXiv (n=2,166; 20%) sharing close to equal representation for the rest of this type of document. A bioRxiv/medRxiv webpage keeping track of medRxiv and bioRxiv’s COVID-19/SARS-CoV-2 preprints index 9,299 manuscripts (respectively; 7,316 and 1,983) as of August 27^th^. Our CORD-19 preprint data (till September 12^th^) for those two sources amounts to 8,289 cases (medRxiv COVID-19, 2020).

**Graph 1:**
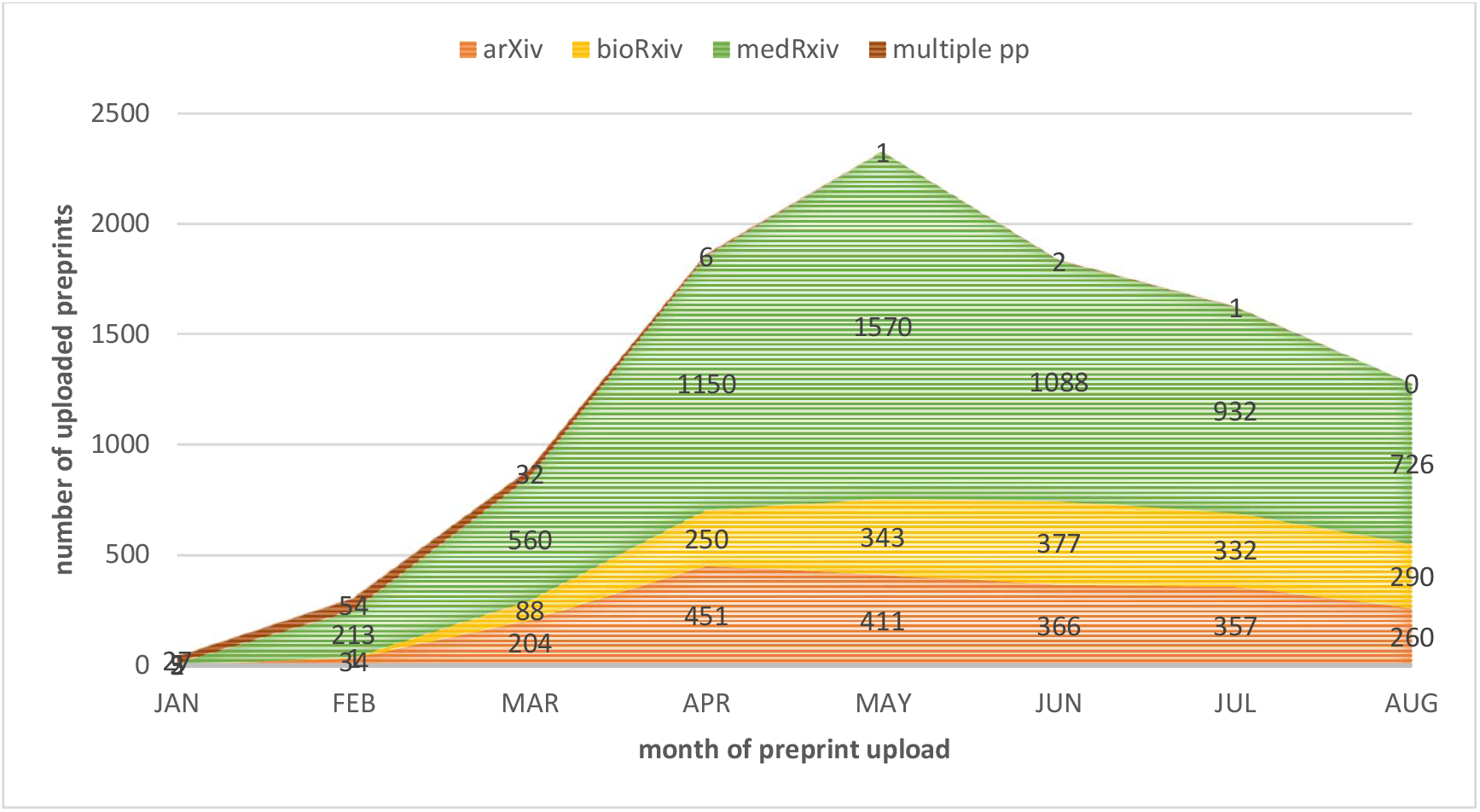
CORD-19 Preprints’ Monthly Upload

### Identification of Published Preprints

Case duplicity in bibliometric datasets is generally quite straightforward to handle, especially when numerical unique identifiers (UIDs) (DOI, PMCID, PUBID, etc) are available. To match preprints to their published counterparts in a giving dataset however is a non-trivial task since UIDs cannot be mobilized. Because of their different institutional origin—online repository vs. academic publisher, preprints and its final published counterparts carry not only distinctive bibliometric metadata (e.g., publication time, title), but also distinctive UIDs.

### Classification Model: Conditional Fuzzy Logic

Giving the unavailability of UIDs to link preprint manuscripts to its published counterparts, or rather to determine if the “conversion event” has occurred, researchers often turn to string-type metadata such as the article’s title instead as a unique id to perform fuzzy matching test. Fuzzy logic is a computing that is based on degrees of truth. In this form of many-valued logic, the result range between 1 (completely true) and 0 (completely false). When comparing two article’s titles using fuzzy logic, we are asking the question “how similar are titles A and B?” Boolean logic, on the other hand, ask “are titles A and B exactly the same?”. Fuzzy string matching, in other words, is the process of finding a string –in our case, a published article’s title—that roughly or approximately match a giving pattern—a preprint’s title. The procedure has several applications such as spell-checking (Pirinen and Linden, 2010), DNA analysis (Fan and Yao, 2011), and spam detection (Singh, Kumari, and Mahajan, 2017).

I use Python package “Fuzzywuzzy” (seatgeek, 2020) that relies on Levenshtein Distance to calculate the differences between sequences and patterns. Lariviere et al. (2014) used fuzzy matching to match a corpus of arXiv preprints with their potential corresponding published versions on Web of Science (WoS). Their method yields an accuracy of 0.50 and an overall F1-score of 0.67. In other words, of all the preprints with a known published version available on WoS, only half were correctly matched using fuzzy logic. Giving this method’s low F1-score, researchers have recently begun experimenting with encouraging results with Bidirectional Encoder Representations from Transformers (BERT; SciBERT), a state-of-the-art NLP tool developed by Google engineers (Devlin, Chang, Lee, and Toutanova, 2018), for that task (Lin et al., 2020).

To address the limitation of fuzzy matching, I developed a conditional approach. In its simple implementation, a preprint’s title is to be compared to each (non-preprint/published) publication’s title from the clean CORD-19 dataset (n=74,153). That is, for each test, the computer program returns a fuzzy score between 0 and 1. With this simple design, the program would compute 74, 153 fuzzy tests for each preprint, but only keep the one(s) with the highest score. The programmer then needs to determine a cut-off point, namely, a fuzzy matching score that tends to minimize false positives/negatives and return valid matches. This final step effectively transforms fuzzy matching into a binary classification {match or no match}.

The two main limitations of this simple implementation of fuzzy matching algorithm are (1) errors (type I and II) and (2) computational time. For instance, to generate a score for our 10,454 preprints the Python program needs to compute 775, 195, 462 fuzzy tests—10454 * 74153. The main driver of errors however rests within the indiscriminatory nature of this design. By using every non-preprint title as a comparator or say, potential match, the program runs the risk of misidentifying one as a match—type I error. This is especially true in a context where all titles include or share similar keyword(s)—see Table 1; filter 2—and where all publications pertain to the same topic—COVID-19/SARS-CoV-2.

Conditional fuzzy matching is an effective solution to handle this problem. The algorithmic strategy consists of *gate-keeping* the actual fuzzy matching step in the program by adding several pre-test conditions that not only considerably reduce the number of fuzzy tests performed per preprint but also reduce the level of type I error. All conditional tests rely on metadata available in CORD-19 dataset. Therefore, to be compared to a preprint’s title, each publication title contained in the COVID-19 dataset goes thought a conditional process that effectively weed out unlikely ‘duplicate’ or published counterpart. The screening process is built around eight conditions (see Table 3).

**Table 3:** Conditional Testing for Fuzzy Matching

| # | Preprint-side | Potential Published Article-side |
| --- | --- | --- |
| 1 | title non-missing | title non-missing |
| 2 | NA | journal name non-missing |
| 3 | author(s) non-missing | author(s) non-missing |
| 4 | NA | difference # author(s) < 5 |
| 5 | NA | first pp_side author in pr_side author(s) |
| 6 | NA | difference title length less than 5 words |

**Table 4:**
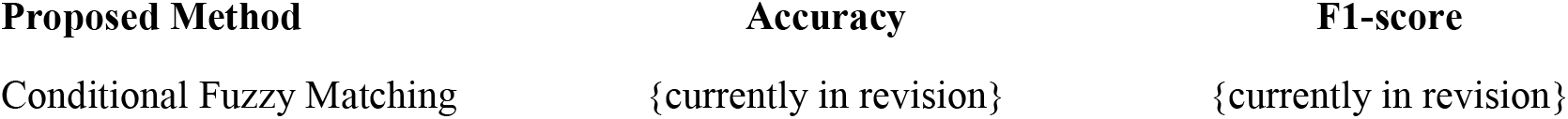
Accuracy and F1-score of Conditional Fuzzy Matching

First, for the Python program to compute a fuzzy test between the titles of a published article and a preprint, the latter needs to have both a non-missing title and non-missing author metadata. Then, the script starts iterating through the list of the 74,153 published articles. But for a fuzzy test to be computed a potential case needs to pass six conditional tests: (1) its title cannot be missing, (2) its journal/venue name cannot be missing, (3) its author(s) listing cannot be missing, (4) the absolute difference between the total number of author(s) of the case and the preprint cannot be larger than 5, (5) the last name of the first author of the preprint need to be found in the potential author’s preprint’s first author’s last name in the list of author(s), and (6) the absolute difference of titles’ length need to be smaller than 5 words. In appendix D, I explain in detail how the algorithm converts this fuzzy matching into a binary classification.

## Results

The CORD-19 dataset I use for my analysis (update 49; September 12th, 2020 version) contains 10,454 unique preprint cases. Giving the medical nature of the topic at hand (infectious disease) it is not surprising to find that medRxiv accounts for nearly two-thirds (n=6,540; 62.5%) of all COVID-19 preprints found in CORD-19 dataset. bioRxiv and arXiv share a similar representation with respectively 17.5% (n=1, 839) and 20% (n=2,166) of the remaining preprint manuscripts. Of all the cases only 123 were uploaded to more than one preprint servers.

To the main research question of this paper, namely, which proportion of COVID-19 preprint manuscripts in CORD-19 dataset were also published in a peer-review journal, the analysis reveals that 19.6% (n=2,049) research documents uploaded on medRxiv, bioRxiv, and arXiv servers between January and early September, 2020 were published as a peer-reviewed article (see Graph 3 for monthly breakdown). Although the method section exclusively focuses on the string fuzzy matching method, in practice I use five different strategies to identify published preprint articles (see Table 5). I call the first two methods – Boolean and fuzzy matching— *internal* methods since they rely exclusively on CORD-19 metadata to identify the preprint corpus’s published counterpart. I call the last three approaches *external* methods since they rely on APIs and web scraping to confirm a preprint’s publication status. The internal methods account for 90% of all linked preprints while the external methods account for 10%.

**Table 5:** Result of Preprint-to-Published-Version Mapping

| Source Preprint | Internal Methods |  |  |  | External Methods |  |  |  |  |  | Published Preprint |  |
| --- | --- | --- | --- | --- | --- | --- | --- | --- | --- | --- | --- | --- |
|  | Boolean |  | Fuzzy Logic |  | bio/medRxiv API |  | arXiv API |  | PMC Scrape |  | source breakdown |  |
|  | n | % | n | % | n | % | n | % | n | % | n | % |
| bioRxiv | 176 | 0.23 | 237 | 0.22 | 36 | 0.23 | NA | NA | 8 | 0.40 | 457 | 0.22 |
| medRxiv | 367 | 0.48 | 772 | 0.71 | 114 | 0.77 | NA | NA | 12 | 0.60 | 1265 | 0.62 |
| arXiv | 221 | 0.29 | 84 | 0.08 | NA | NA | 27 | 1 | 0 | 0 | 332 | 0.16 |
| **n** | 764 | - | 1093 | - | 150 | - | 27 | - | 20 | - | **2054** | - |
| **%** | 0.37 | 1 | 0.53 | 1 | 0.07 | 1 | 0.01 | 1 | 0.01 | 1 | **0.20** | 1 |

**Graph X:**
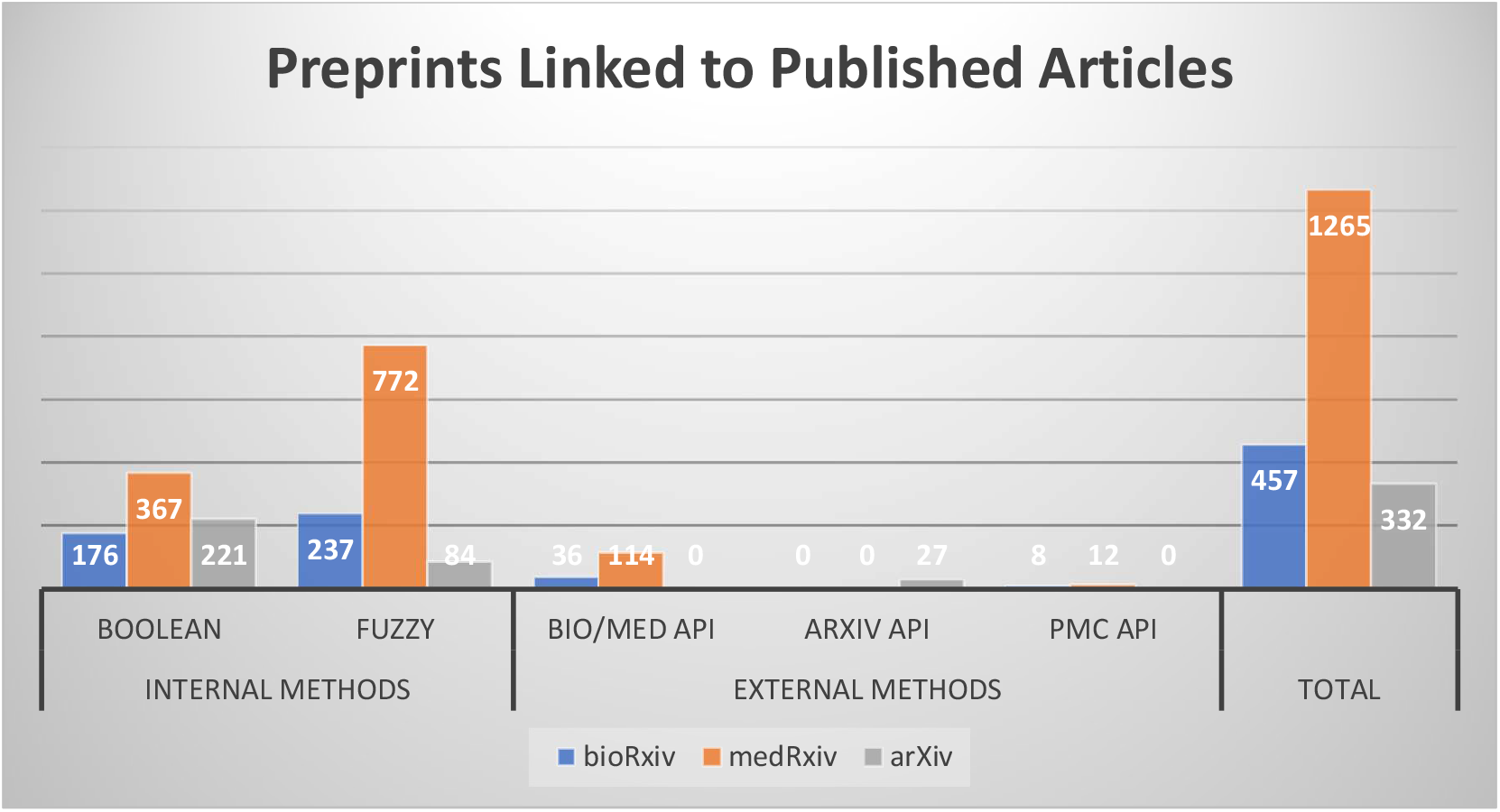
Clustered Column Chart of Preprints Linked to Published Articles

### Internal Methods: Conditional Usage of CORD-19 Metadata

First, running a Boolean test on each preprint title, I identify 764 cases as published preprint articles. In other words, I found 7.3% of preprints for which the title was exactly the same as its peer-reviewed counterpart’. In a second time, I ran a conditional fuzzy logic script on all remaining cases that returned a negative or false Boolean result (n=9,690). The Python program yields a final list of 1093 matched cases, or 10.6% of all CORD-19 preprints. That is, in addition to the Boolean matches, the algorithm identified 1093 peer-reviewed articles as the published counterpart of a COVID-19 preprint.

Of those, 365 (33%) were matched with a fuzzy score of 100 (see Appendix B for distribution of fuzzy matching results). One might ask why the previous Boolean test did not return a True result for those very cases. It has to do with the way Python’s fuzzy matching package prepare strings before computing a test. In other words, for those 220 cases, the only difference was a period (‘.’) at the end of the title. With the fuzzy matching algorithm, those symbols are automatically removed from each string before a test while non-alphabetic symbols remained while computing a Boolean test. If fuzzy string logic appears to be efficient or accurate in identifying preprints’ published counterparts, why implementing a Boolean test as a first step? The reason has to do with computational time, namely, reducing the number of preprint cases needed to be processed for the fuzzy test.

### External Methods: Bibliometric APIs as Additional Resources to Identify Published Preprints

In addition to using CORD-19 metadata and fuzzy logic to identify preprint manuscripts that have a published counterpart, I also use freely available bibliometric APIs to detect more cases harboring a publishing status. Some digital repositories have record-linkage functionality that automatically updated its preprint metadata to reflect its publication history—e.g., journal, title, date, etc. Preprint servers also encourage authors to update this information themselves for their accepted manuscripts (Lin et al., 2020). By querying bio/medRxiv and arXiv APIs as well as scraping PMC preprint webpages, this second effort produces 192 new cases.

#### bioRxiv API

When querying bioRxiv API with the list of all remaining ‘unmatched’ CORD-19 preprints originating from that digital repository, I found a total of 146 preprints that had a DOI generated by a peer-reviewed journal which is a reliable indicator of a published status.

#### PMC Web Scraping

For unmatched CORD-19 preprint cases with a PMC id and/or a PUBMED id, I query the PubMed Central website using Python BeautifulSoup and requests packages to determine if they were linked to a peer-reviewed version as well. The string occurrence of “Update of” or “This article had been published” is a reliable indication that the preprint was published in a peer-reviewed journal. This method produced 19 new cases.

#### arXiv

Finally, I use the Python wrapper ‘arxiv’ to access arXiv API. Compare to other well-established preprint servers, arXiv has the particularity of not automatically generating a DOI for manuscripts uploaded on its platform. That said, arXiv “cooperates with Inspire (formerly SPIRES) to provide an automatic update to DOI information and journal references if a preprint is published (Lin et al., 560).” This last approach allowed me to further identify 27 additional arXiv manuscripts related to a peer-reviewed version.

**Graph 2:**
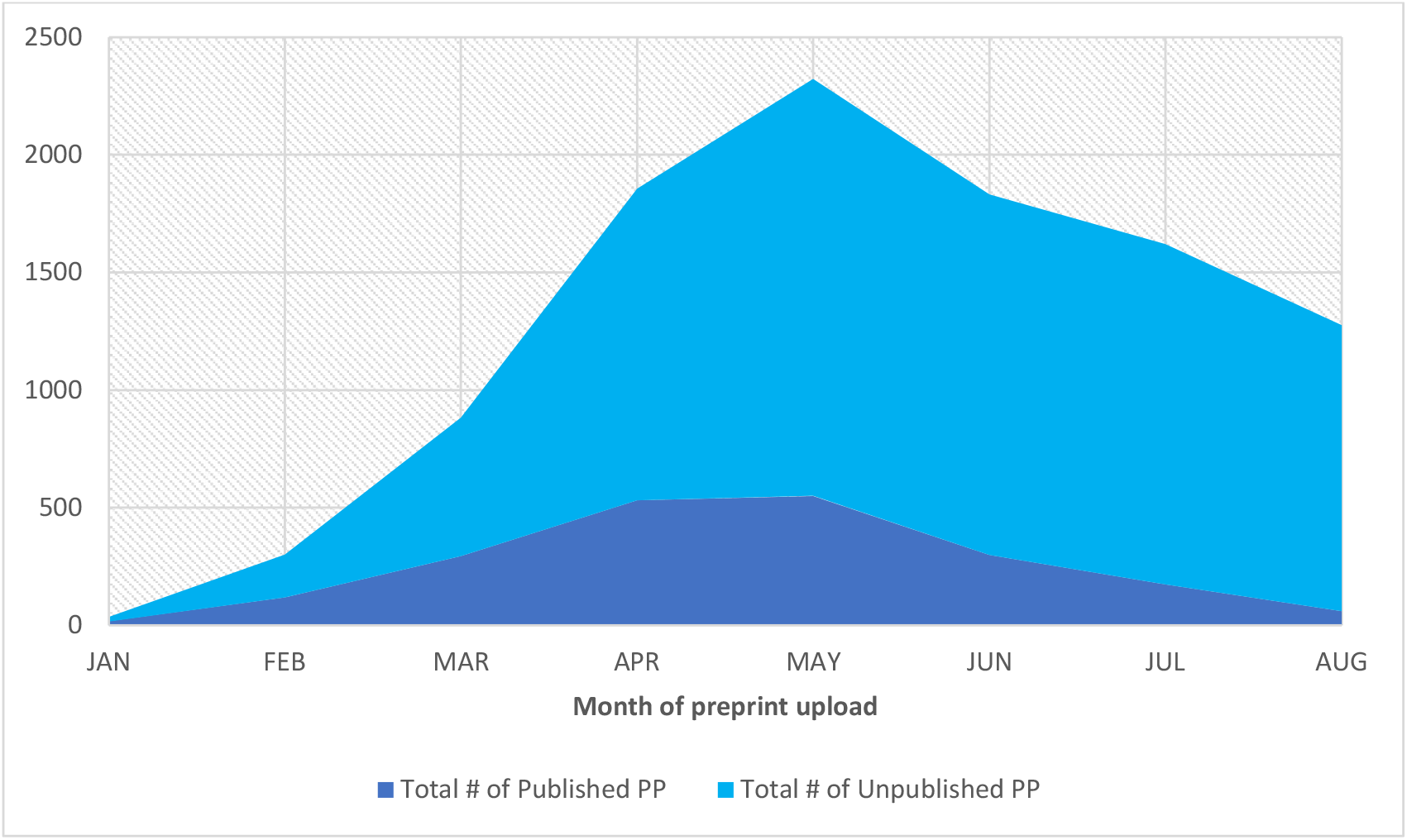
Stack Chart – Monthly Distribution of Preprint’s Publication Status (raw)

**Graph 3:**
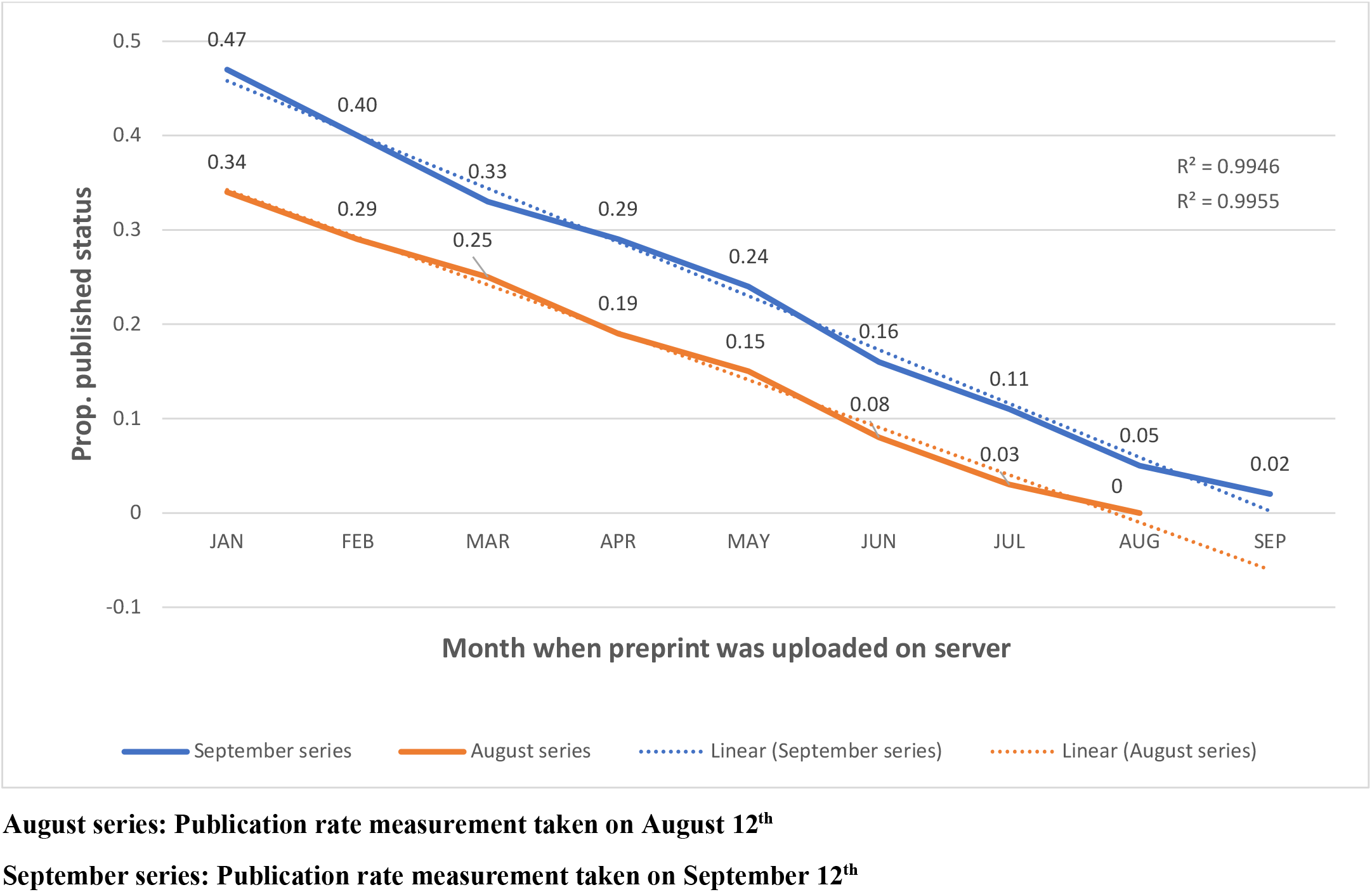
Monthly Distribution of Preprint’s Publication Status (proportion + linear trendline)

**Graph 4:**
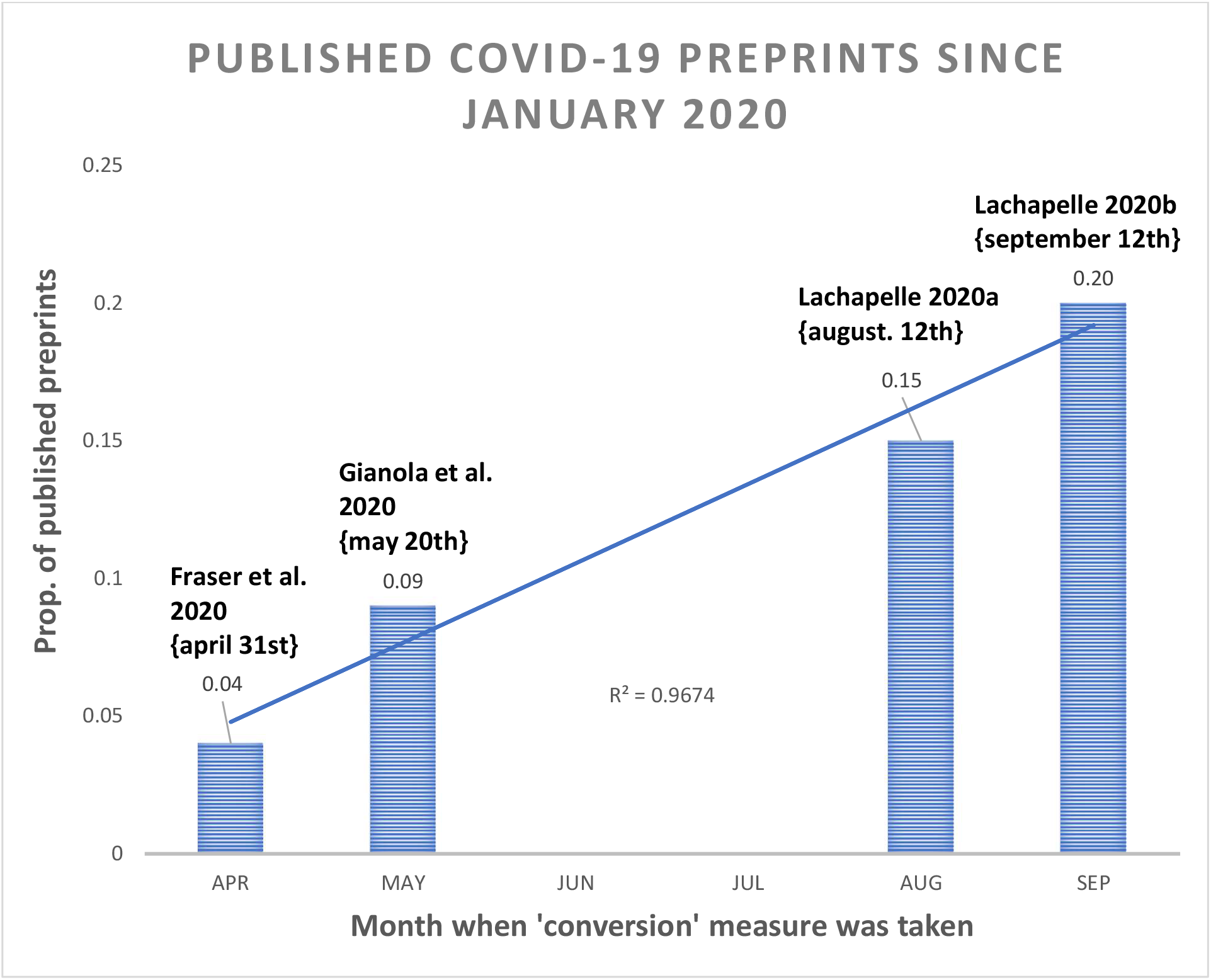
Bart Chart – Four Measurements of COVID-19 Preprints’ Conversion Rate (proportion + linear trendline)

### Introducing a new API: Upload-or-Publish

The landscape of scientific communication has shifted drastically since the beginning of the current SARS-CoV-2 pandemic. As mentioned in the introduction, several initiatives have sprung since March to generate stronger connective tissues between the preprint infrastructure and the traditional peer-review pipeline. Biomedical bibliometric powerhouses like PubMed and PMC have started indexing preprints on their own platforms. What is missing however is a robust tool that tracks changes in the publication status of preprint papers.

I introduce Upload-or-Publish {UoP}, a micro-API to query COVID-19 preprints’ publication status. The micro-API service hosted on PythonAnywhere offers a free, programmatic way for a client to make a query on a COVID-19 preprint’s (or a batch of COVID-19 preprints) publication status. The API returns a JSON dictionary with CORD-19’s enhanced metadata. If the preprint’s publication status is positive, then, the API returns a JSON containing metadata of the published article. The UoP API, at least in its current implementation, requires no keys/authentication.

### Institutional Users

I developed the API with mainly two groups of users in mind: institutional clients and bibliometric researchers. In their efforts to deal with the growing position of preprints in the making of science, I strongly believe bio/medRxiv repositories curators, leading biomedical bibliometric platforms—PMC, PubMed, as well as dominant proprietary operators ranging from academic journal publishers to scientific indexing services are in a serious need of fine-grain metadata when it comes to preprints’ publication status. In the past several months we witnessed several initiatives that strive to integrate and recognize the key role preprints can play in the scientific ecosystem, but none have developed an information system to track preprints’ publication status. This is what UoP proposes.

### Preprint Servers Coverage

The current version (beta) covers the three main preprint servers included in the CORD-19 data project: arXiv, bioRxiv, and medRxiv. As we know, there are dozens upon dozens of online English-language preprint repositories. In the first survey of preprint servers in the context of the current pandemic, Fraser et al. (2020) reported that “eleven of the 31 preprint servers included in our dataset hosted over 100 COVID-19 related preprints each” (lines 126-27). Therefore, one of my priorities is to extend UoP’s repo coverage. NIH iSearch COVID-19 publication database is a great first step in that direction since it includes three preprint servers not systematically covered by the CORD-19 dataset, namely, ChemRxiv, SSRN, and ResearchSquare.

## Discussion

My analysis reveals that about 20% of COVID-19 preprint manuscripts in the CORD-19 dataset uploaded on three major repository servers between January and early September 2020 were published in peer-reviewed venues. This latest measurement represents a 5% increase for the month of August (Graph 4). When compare to Gianola et al.’s conversion rate of 8.6% for preprints uploaded between January and May, 2020, the result suggests more than a two-fold increase over the last four months (Graph 4). We can compare our measure to Gianola et al.’s finding without arXiv preprints since they did not include that server in their purposive sample of preprint repositories. Going so, our conversion rate increased by nearly 1% to reach 21% (not indicated in Graph 4).

So, why is the proportion of COVID-19 published preprints so low, and why are we witnessing an upward trend? Regarding the former question, Gianolo et al. (2020: 16) speculate that time is a key variable to consider. In other words, converting a preprint manuscript into a published peer-reviewed takes time which is why the scientific community turned to preprint repositories as a means of research dissemination and communication early on in its effort to study the pandemic. Preliminary results (not shown), using CORD-1 metadata, indicate an average length of 60 days for a preprint to get published. A visualization of preprints’ publication rate using a breakdown *per month of server upload*, indeed, reveals that the overall figure of 20% can mask the core temporal dynamic at play.

The results from Graph 3 showcase how time plays a key role in the process of transforming a preprint manuscript into a published peer-reviewed article. The near-perfect correlation between the linear trendline and the proportion of published preprints grouped by month of server upload (r=0.99) for both the August and the September series capture a positive relationship between the time elapsed since the first server upload of preprint and a preprint harboring a published status. For instance, as of mid-September, close to 50% of preprints uploaded in January were published.

That said, there are good reasons to predict that as time goes by this positive linear relationship between time and publication status will take a right-skewed shape. In a pre- and non-COVID-19 related-context, for bioRxiv, findings indicate 75% of published preprints appear in a journal within eight months of their initial upload (Abdill et al., 2019a: 27; similar results – Iwema et al., 2016). For arXiv, this figure is 12 months (Lariviere et al., 2014). Another research on bioRxiv preprints found a probability of 48% for a successful conversion from a preprint to the published article within the first six months of being uploaded (Serghiou and Ioannidis, 2018). In our case, the six moths mark—CORD-19 preprints uploaded in March— recorded a conversion rate of 33% (Graph 3). It took eight months for the CORD-19 preprints uploaded in January to reach a published rate of 47%.

When compare to Lin et al. (2020) publishing rate of 77% for arXiv repository’s computer science-related preprints uploaded between 2008 and 2017, then, our finding for COVID-19 preprints does appear to be on the lower side of the conversion spectrum. As reviewed in the introduction, measures of publication rate for bioRxiv and arXiv preprints indicates that most manuscripts eventually get published (Abdill et al., 2019; Lariviere et al., 2014; Schloss, 2017). The author knows of no comparable research for medRxiv preprints. In the rest of the discussion, I develop on three potential reasons why it is very unlikely that the overall proportion of published COVID-19 preprints will reach a publication rate higher than 50%.

### Competition and Epistemic Oversupply

Let me now considerer another likely contributing factor for the relatively low publishing rate of COVID-19 preprints, which I call ‘epistemic oversupply’. I mean by that the fact that giving the high level of knowledge production, scientific work on COVID-19, including preprint formats, run the risk of suffering from accelerated obsoleteness as several teams around the globe work, submit, and try to publish research on the same topic independently of each other. After all, uncertainty is a key feature of scientific labor (Whitley, 1984). Scientific activities rely on cooperation to advance towards its goal, but like any other human pursuits, competition also plays a vital role in the way scientific fields operate (Merton, 1973; Bourdieu, 1984). As countries (Keck, 2020; Horowitz, 2020) and pharmaceutic giants (McDonnell, 2020; Callaway, 2020) are competing in the billion-dollar vaccine race against COVID-19, teams of scientists are also competing for symbolic and material capital—e.g. huge grants and funding—as they rush to produce novel insights on the deadly virus. Some scholars have framed this problem of competition in the context of COVID-19 as one of waste (Casigliani et al, 2020; Gale, 2020). “Some replication of studies is important, but unnecessary duplication of studies is wasteful” (Glasziou, Sanders, and Hoffman, 2020).

Therefore, in a context of high competition, it is possible that behind a low conversion rate hides a high rejection rate. It might not simply be that authors of preprints are not submitting to peer-reviewed journals, but rather that they were rejected because of “unnecessary duplication”. Or, in the face of discovering too many similar studies, researchers decide to abandon or rework the focus of their work. A social survey of contributors to COVID-19 preprints would be an appropriate study design to collect data on mechanisms, or emerging practices around the “after-life” of preprint manuscripts.

### Performative Assent – Peer-Skippers and The New Distributed Nature of Peer-Review

There are ample shreds of evidence to suggest that in the context of “the lightning-quick progression of a pandemic” (Fraser et al., 2020: line 395), a fair share of scientists see the traditional system as an unfit channel of knowledge dissemination and preprint servers as an effective alternative for sharing research. As Thomas Kuhn, the author of *The Structure of Scientific Revolutions* (1962) would remind us, with its ongoing effort to study and respond to the global pandemic, the scientific community left ‘normal science’ to enter an “extraordinary” phase of development. One might be tempted to brush off the notion that a scientific revolution is underway, but *an extensive revision to existing scientific practices—*such as what we are witnessing with the key role of preprints in helping maintain an effective research ecosystem— is a core characteristic of a scientific revolution. As we know, extraordinary phases in the history of science are marked by a sharp acceleration of progress. Covid-19 research represents one of the most intense scientific efforts in the modern history of science (Vlasschaert, Topf, Hiremath, 2020). Scientists and journal editors and reviewers alike were (and arguably still are) overwhelmed with a tsunami of COVID-19 papers (Brainard, 2020; Kwon, 2020). And this extreme phase of knowledge production on the virus is not behind us. The week of August 24^th^ replaced the week of May 11^th^ (2,585 pubs) for the period recording the most COVID-19 publications indexed by PubMed in a single week with 3,241 (LitCovid, 2020).

Times of crisis tend to see the emergence of new forms of legitimate knowledge. As explained by the sociologist of science Steven Epstein in his seminal book *Impure Science* (1996), during the HIV pandemic, not only did AIDS activists notoriously confront medical authority, but they also durably helped transformed deep-rooted research practices. For instance, at the time high-status medical journals vigorously forbade the early release of scientific research before publication. But with the pressure of AIDS activists, early release emerged as an acceptable practice. Early release is now a SOP in health-crisis like the current COVID-19 pandemic. Certainly, the case of the preprint is different than early release in that the latter has already been peer-reviewed while the former remains in a liminal state; a scientific research stuck between high visibility and questionable legitimacy. As surveyed in the introduction, this very characteristic is what makes preprints a very contentious form of scientific knowledge.

But although this line of argument can explain the popularity of preprint servers, it does not totally explain preprints’ low conversion rate. Put differently, what interests me as a sociologist of science is the question of why researchers might decide not to submit their preprints for peer-review—we might want to call them the *peer-skippers*. Granted, to empirically test how much this decision help account for preprint’s low conversion rate, we need survey data on submission and rejection of preprints. But it is still worth asking how *peer-skipper* researchers deal with/justified the fact that their preprint work will not receive the ‘traditional’ stamp of certification. They might not be alone. In recent years, some preprint advocates began breaking rank with the established standard operating procedures as they argue preprint research can receive scientific assent outside the traditional peer-review process (Singh Chawla, 2017; Kaiser, 2017). Those defending the idea of final version preprint (or *peer-skipping*) use the distinction between ‘pre-publication peer-review’ (pre-ppr) and ‘post-publication peer-review’ (post-ppr) to make their point. From this perspective, the main difference between a peer-reviewed article and a preprint is that the former received both pre- and post-ppr while the latter only received post-ppr. But, against the belief that preprint servers are a digital far-west where anything can be uploaded, a recent analysis of 44 preprint servers’ uploading procedures found that the majority have quality-control mechanisms in place and 32% include researchers in the vetting process (Kirkham et al., 2020). Although not to be mistaken for a formal pre-ppr, it does indicate that most biomedical preprints vet preprints for quality-control before officially making manuscripts available on their repositories. Finally, preprint advocates point to the effectiveness of post-ppr, or ‘post-upload’ peer-review process. After all, the retraction of both preprints and peer-review articles during the pandemic usually happens because of post-publication quality control by fellow-researchers using, interacting, and ultimately evaluating scientific works (Singh, 2017; Yeo-Teh and Tang, 2020; RetractionWatch, 2020). A scholar even suggested that the peer-review process during the pandemic might be, in some cases, even more “imperfect” or “porous” than usual, that is, “non-existential” (da Silva, 2020: 1). In the medial field, the journal Cureus used since 2009 crowdsourcing for its peer-review process as well as both pre- and post-publication peer-review.

I am not suggesting those ‘fringe’ voices advocating for the preprint culture as an alternative system has spread their gospel during the pandemic—although it might be the case—but I believe one of the contributing factors to the low conversion rate of COVID-19 preprints is linked to this new form of post-ppr. Da Silva (2020: 1) call it “open public scrutiny for preprints”. That is, even though preprints are not formally reviewed before being uploaded to preprint servers, they are often commented on and shared on social media—including blog posts (Vlasschaert, Topf, Hiremath, 2020)—by laymen and experts alike (Majumder et al, 2020; Fraser et al., 2020), downloaded, and periodically updated by its author(s) once online, and cited – even more cited than published articles (Gianola et al., 2020). Some independent initiatives have also “arisen whereby reviews are posted in the comments section of preprint servers and hosted on independent websites” (Fraser et al.; from 34). It is not unlikely that author(s) perceived these social processes as something I call “performative assent”, a type of approving green light of sort that differs from the traditional pre-publication review. Preprint servers are not only unusually fast in sharing research, but their public nature might increase the scrutiny scientific manuscripts usually received. This feature may be preprint servers’ best protection against bad science. A poor manuscript is not about to receive the private feedback of a few peers, but potentially the critical gaze of hundreds or thousands of readers on several platforms. More scrutiny prevents bad uploads. Is this form of distributed review or watchfulness as sociologist Keith Hampton called it (2015) a closer implementation or realization of open science? This might help explain how research on the textual similarity of 12, 202 scholarly preprints and their final published counterparts in STEM disciplines found no significant difference between preprints and published versions (Klein et al., 2019). Similar analyses by Fraser et. al (2020) reveals that only 4.9% of COVID-19 published preprints “display[ed] major changes in the conclusions”.

The concept of *performative assent* captures the notion that preprints can gain a legitimated status as users evaluate and interact with the document. Could it be that performative assent disincentivize author(s) to carry their research through the classic channel of peer-review because they already received a level of approval based on the public and expert interaction with their text, especially in a digital world where all those metrics (views, download, citation, alt-metric, etc..) are a click away. Findings indicate that compare to non-COVID-19 preprints, COVID-19 preprints received 15 times more views and 30 times more downloads (Fraser et al., 2020).

### Limitations & Future Directions

In this article, the main goal was to measure the proportion of COVID-19 preprints that were published in peer-reviewed journals between January and August 2020. I followed with a discussion, although perhaps speculative, on some reasons that can help explain the low publication rate of COVID-19 preprints. More notably, I coined the concept of “performative assent” to articulate how a sets of relational metrics – view, download, citation, peer-comment, altmetric – can, in the age of social media, help construct scientific legitimacy (or the illusion of) in a loosely-defined post-publication peer-review process. I also point that in a context where multi-level cooperation (civil society, government, private sector, science) is paramount to fight the pandemic, scientific competition remains well and alive. In other words, redundancy happens as researchers fight for the limited prime space in top academic journals. I encourage students of preprint culture and knowledge production to use social survey to start documenting the experience and decision-making of COVID-19 researchers who are users of preprints’ digital repositories. Questions pertaining to their experiences of submission to peer-reviewed venues, rejection, and revision—the whole cycle—can help illuminate emerging practices and tactics around preprints. Of particular interest is researchers’ thinking (and justification) around the decision of not submitting their manuscript to peer-reviewed journals—peer-skippers.

My next priority is to empirically test my performative assent theory. The rather counter-intuitive hypothesis posits that the more a preprint manuscript received attention—tweet, comments, download, etc—the less it is likely to be submitted for formal peer-review. Since Rxivist API allows the retrieval of preprints’ twitter metadata and Disqus API allows the retrieval of bioRxiv and medRxiv’s comments on its preprints (for example, see Fraser et al., 2020), I am very confident I can test this important theory.

That said, more quantitatively-inclined scholars will also want to use zero-inflated negative binomial regression analysis to model factors predicting the likelihood for a preprint to get published. In their recent work, Lin et al. (2020:568) identified version history, number of authors, article length, number of references, citation count, number of figures and tables as potential variables to include in predictive models. That said, researchers will need to mitigate the influence of important confounding factors, more notably the existence of platforms like Nature’s Outbreak Science Rapid PREreview and MIT’s Rapid Review (RR:C19) which can select preprints for a round of peer-reviewing. Time is also a critical dimension to account for. Time-lagged or survival analysis are idle tools to model the temporal component of publishing.

In terms of the author’s own plan, the first task is to merge NIH iSearch COVID-19 publication database’s preprint articles to CORD-19 dataset as it will add three additional preprint servers (ChemRxiv, SSRN, and ResearchSquare) to the list of online repositories curating COVID-19 scientific content. An important data science contribution would be to start integrating COVID-19 preprints from all major English-language preprints servers. I am also planning for the next iteration of this project to provide more granularity in the analysis and exclude cases where the peer-reviewed version is published before the date of the preprint’s uploading.

The pandemic is unquestionably a stress-test for the preprint infrastructure and the traditional system responded with a host of initiatives to normalize the peer-review process of preprints. On their side, online repositories of scientific preprints will need to standardize their data-management practices. The establishment of the Upload-or-Publish API pipeline and the systematic tracking and updating of preprints’ publication status is an important step. Beyond UoP micro-API, it would also not be surprising to see third-party initiatives like Rxivist (Abdill and Blekhman, 2019b) or others working towards the consolidation of all English-language preprint curation services data into one platform or API service. But until then, string matching techniques either using fuzzy matching or state-of-the-art AI will remain our best tools to further document the changing relationship between public drafts of scientific work and their finalized legitimate versions.

### Conclusion: From Publish or Perish to Upload or Perish?

The pandemic will leave deep scars of change on most human institutions. The uneasy relationship between preprint servers and the traditional system of peer-review is one such example. In a post-pandemic scientific landscape, some will return to their lab converted to the motto of “upload or perish” where preprints will stand at the center of their cultural practice of knowledge production. Some will have their preprints picked by AI algorithms, then reviewed and published regardless of their preferences. Some will have just discovered online repositories for their work. Some will make strategic usage of preprint servers only for certain types of contributions.

In other words, we likely see a wider acceptance of the preprint online repositories as a ‘parallel’ system – a challenger to the traditional system of science. Underlying this potential radical change is the very way academic work can gain scientific assent, from a close to an open system. Only time will tell if the future of science will be done by uploading one preprint at the time, or still by submitting one manuscript at the time.

## Data Availability

I use Covid-19 Open Research Dataset (CORD-19) to calculate COVID-19 preprint corpus' conversion rate to peer-reviewed articles. Arguably the most ambitious bibliometric COVID-19 project, CORD-19 is the collaborative effort between the Allen Institute for AI and half a dozen organizations including NIH and the White House (for more details, see Wang et al., 2020). This is an open-source dataset.
I also used bioRxiv API pipeline to determine if COVID-19 preprints were associated with a peer-review final counterpart.
I also scraped pubmed and pmc NIH's websites for the same purpose.
Finally, I use the Python 'wrapper' package "arxiv" to query arXiv aPI to, again, determine if certain COVID-19 arXiv preprints had also been a published peer-reviewed journal.

https://www.kaggle.com/allen-institute-for-ai/CORD-19-research-challenge

https://api.biorxiv.org/details/biorxiv/

https://pubmed.ncbi.nlm.nih.gov/

https://www.ncbi.nlm.nih.gov/pmc/articles/

https://github.com/titipata/arxivpy

## Appendix A Estimation of Ranges for True Publication Rate

{currently being revised after receiving post-upload peer-review feedback}

## Appendix B Fuzzy Logic Results

**Table 11:** Fuzzy Logic Results

| **Preprint Source -->** | **arXiv** |  | **bioRxiv** |  | **medRxiv** |  | **Total** |  |
| --- | --- | --- | --- | --- | --- | --- | --- | --- |
| **Fuzzy Score** | **n** | **%** | **n** | **%** | **n** | **%** | **n** | **%** |
| **100** | 45 | 0.54 | 85 | 0.36 | 235 | 0.30 | **365** | 0.33 |
| **91 - 99** | 16 | 0.21 | 79 | 0.33 | 236 | 0.31 | **331** | 0.30 |
| **81 - 90** | 4 | 0.05 | 37 | 0.16 | 144 | 0.19 | **185** | 0.17 |
| **71 - 80** | 13 | 0.15 | 28 | 0.12 | 120 | 0.16 | **161** | 0.15 |
| **61 - 70** | 2 | 0.04 | 7 | 0.03 | 25 | 0.03 | **34** | 0.03 |
| **51 - 60** | 4 | 0.05 | 1 | 0 | 12 | 0.02 | **17** | 0.02 |
| **41 - 50** | 0 | 0 | 0 | 0 | 0 | 0 | **0** | 0 |
| **31 - 40** | 0 | 0 | 0 | 0 | 0 | 0 | **0** | 0 |
| **21 - 30** | 0 | 0 | 0 | 0 | 0 | 0 | **0** | 0 |
| **under 20** | 0 | 0 | 0 | 0 | 0 | 0 | **0** | 0 |
| **Total** | **84** | **1** | **237** | **1** | **772** | **1** | **1093** | **1** |

## Appendix C How to Use the UoP API {beta}

The current beta-version only allows one route “http://heibufan.pythonanywhere.com/json/pp_meta/doi”.

For example, if a client wants to determine the publication status of a specific COVID-19 preprint, using its DOI, the URL should be:

➢ http://heibufan.pythonanywhere.com/json/pp_meta/10.1101/2020.03.19.998179

The returned JSON will look like this:

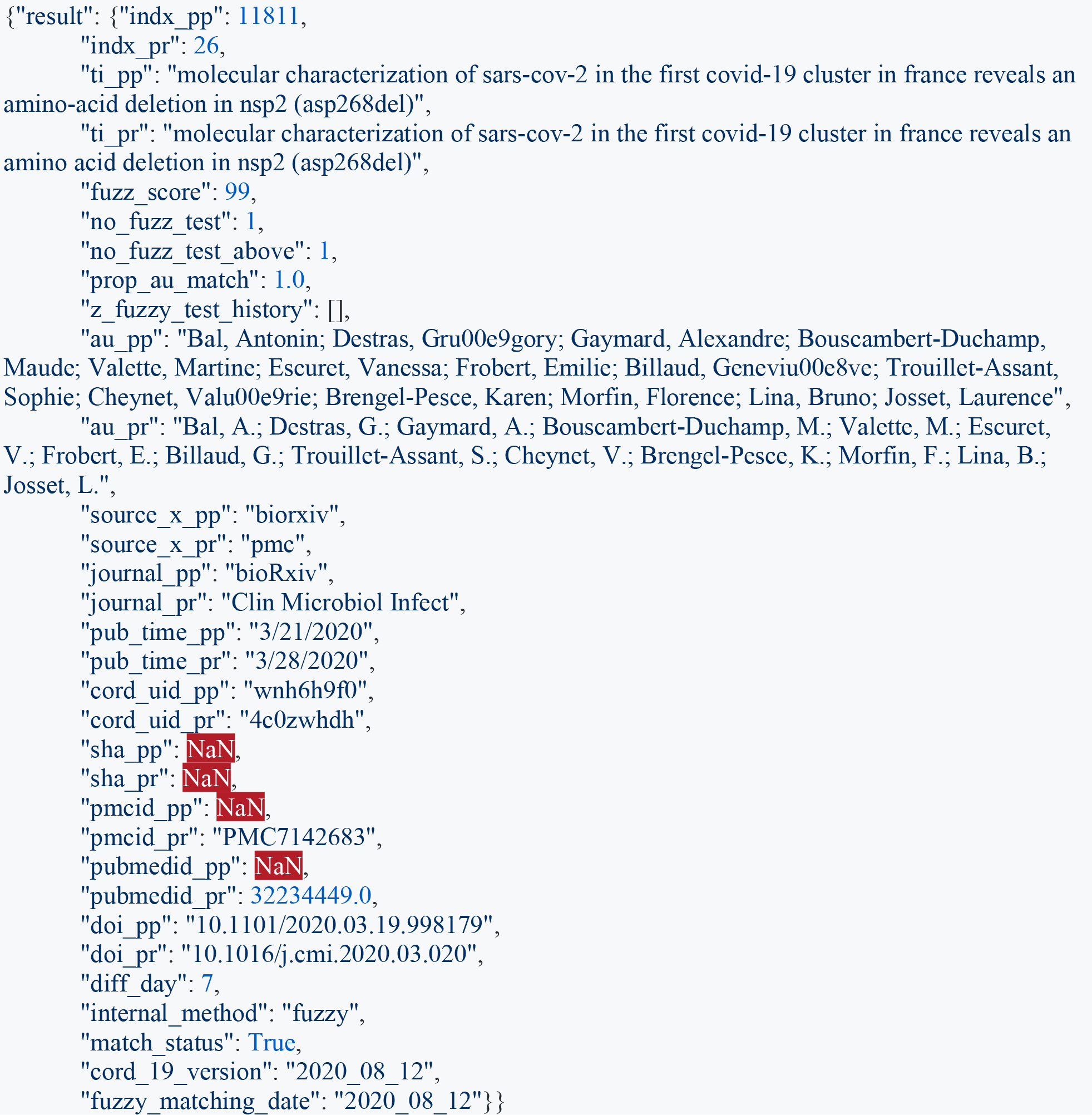

### UoP Documentation

When parsing the returned JSON for a specific preprint query, please note the most important returned metadata is *match_status* <u>(#30 in Table 12)</u>:

**Table 12:** UoP Documentation Description

| # | List of Abbreviations Used in Documentation |
| --- | --- |
| 1 | "pp" stands for preprint manuscript |
| 2 | "pr" stands for peer-reviewed article |
| 3 | "(c)" indicates CORD-19 metadata |

✓ A Boolean value of True indicates that the preprint was linked to a peer-reviewed published counterpart
✓ A Boolean value of False indicates that the preprint was NOT linked to a peer-reviewed counterpart.
✓ A preprint returned a False value for match_status should NOT be considered to have been published in a peer-reviewed journal. Only a preprint that returned a True value for match_status can be considered to have been published in a peer-reviewed journal

| # | Metadata | Description |
| --- | --- | --- |
| 1 | 'indx_pp' | internal working id for developer |
| 2 | 'indx_pr' | internal working id for developer |
| 3 | 'ti_pp' | title of preprint (c) |
| 4 | 'ti_pr' | title of published/peer-reviewed article linked to preprint (c) |
| 5 | 'fuzz_score' | fuzzy logic score obtained when ti_pp and ti_pr were compared |
| 6 | 'no_fuzz_test' | total number (raw) of fuzzy logic score(s) computed (see methods) |
| 7 | 'no_fuzz_test_above' | total number of fuzzy score(s) computed that were above the cut-off point of 0.50 |
| 8 | prop_au_match' | proportion of pp's authors' last names that was found in the list of authors of the pr article |
| 9 | 'z_fuzzy_test_history' | list/array of results of all fuzzy matching tests computed with score >=50 |
| 10 | 'au_pp' | list of authors (preprint) (c) |
| 11 | 'au_pr' | list of authors (published version) (c) |
| 12 | 'source_x_pp' | bibliometric source where CORD-19 retrieved metadata from (c) |
| 13 | 'source_x_pr' | *ibid* |
| 14 | 'journal_pp' | online preprint repository where manuscript was uploaded (c) |
| 15 | 'journal_pr' | peer-review journal venue where preprint was submitted and published (c) |
| 16 | 'pub_time_pp' | date of preprint upload (c); note: a preprint can have multiple uploaded versions. Still need to validate that CORD-19 consistently use v1 date |
| 17 | 'pub_time_pr' | date of peer-reviewed article's publication (c); note: still need to validate that CORD-19 use date of "e-print" which is the date when the article is made available online. |
| 18 | 'cord_uid_pp' | preprint's id (c) - note: CORD-19 unique id |
| 19 | 'cord_uid_pr' | *ibid* |
| 20 | 'sha_pp' | id and name of the full-text pdf parsed in a JSON format (c); a sha_id indicates that for that article CORD-19 has not only the full text in PDF format available for download, but also that CORD-19 used the GROBID tool to fully parse the PDF in JSON and/or XML format. |
| 21 | 'sha_pr' | *ibid* |
| 22 | 'pmcid_pp' | pub med central id (c) |
| 23 | 'pmcid_pr' | *ibid* |
| 24 | 'pubmedid_pp' | pub med id (c) |
| 25 | 'pubmedid_pr' | *ibid* |
| 26 | 'doi_pp' | digital unique identifier (c); note: arXiv does not automatically generate DOI for the preprint manuscripts its curated. Therefore, an arXiv preprint with a DOI indicates one of two possible scenarios-(1) preprint was also uploaded on another server; (2) preprint has a published counterpart and DOI was generated by an academic publisher/journal. |
| 27 | 'doi_pr' | *ibid* |
| 28 | 'diff_day' | difference in day between pp upload and pr publication; note: see 'pub_time_pp' |
| 29 | 'internal_method' | record-linkage method used to match pp and pr; Bool or Fuzzy |
| 30 | 'match_status' | True: preprint has a peer-reviewed counterpart, False: preprint has no peer-reviewed counterpart. This is the most important metadata to consider. Preprint with a match_status of False should NOT be considered as having been published in a peer-reviewed journal. |
| 31 | 'cord_19_version' | version of CORD-19 dataset used to generate UoP record-linkage API |
| 32 | 'fuzzy_matching_date' | date {year month day} when the record-linkage code was executed |

#### Snippet Code Example in Python

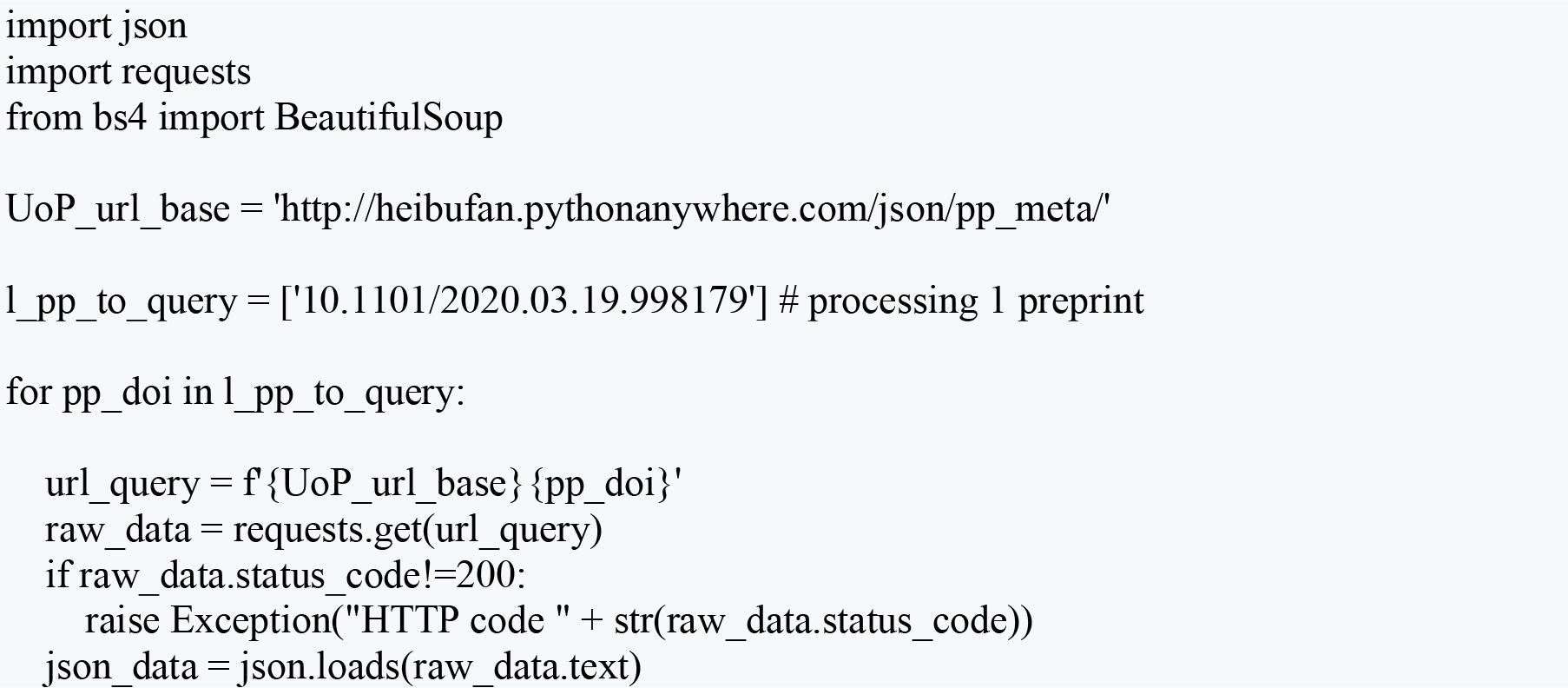

## Appendix D Fuzzy Logic, Cut-Off Points, and Binary Classification

{author’s note: coming in next version}

## Notes

### Competing Interest Statement

The authors have declared no competing interest.

### Funding Statement

The author received no funding for this work.

### Author Declarations

This research is not require any approval or exemption from any IRB/oversight body at my home institution.

### Summary of Updates

full sensitivity analysis in process of revision following post-upload peer-review on Twitter my main finding mentioned in the abstract showcasing that 20% of preprints uploaded between January and September are published in peer-review venues is accurate, but it doesn't highlight the large difference in the publication rates depending on when preprints were uploaded. My findings section in the abstract is now highlighting this relationship.

## References

Nature Bio Editors. (2017). Methods, preprints and papers. Nature Biotechnology.

Nature Bio Editors. (2020). All that’s fit to preprint. Nature Biotechnology.

Abdill, R. J., & Blekhman, R. (2019a). Meta-Research: Tracking the popularity and outcomes of all bioRxiv preprints. Elife, 8, e45133.

Abdill, R. J., & Blekhman, R. (2019b). Rxivist. org: Sorting biology preprints using social media and readership metrics. PLoS biology, 17(5), e3000269.

bioRxiv. (2020). Submission Guide. Retrieved from https://www.biorxiv.org/submit-a-manuscript

Bourdieu, P. (1984). Homo Academicus (Paris: Les Éditions de Minuit, 1984).

Callaway, E. (2020, April 30). Score of coronavirus vaccines are in competition – how will scientists choose the best?. Nature. Retrieved from https://www.nature.com/articles/d41586-020-012472

Casigliani, V., De Nard, F., De Vita, E., Arzilli, G., Grosso, F. M., Quattrone, F., … & Lopalco, P. (2020). Too much information, too little evidence: is waste in research fuelling the covid-19 infodemic?. bmj, 370, m2672.

Chen, Q., Allot, A., & Lu, Z. (2020). Keep up with the latest coronavirus research. Nature, 579(7798), 193–193.

Chiarelli, A., Johnson, R., Pinfield, S., & Richens, E. (2019). Preprints and Scholarly Communication: An Exploratory Qualitative Study of Adoption, Practices, Drivers and Barriers. F1000Research, 8.

Cobb, M. (2017). The prehistory of biology preprints: A forgotten experiment from the 1960s. PLoS biology, 15(11), e2003995.

da Silva, J. A. T. (2020). An alert to COVID-19 literature in predatory publishing venues. The Journal of Academic Librarianship.

Desjardins-Proulx, P., White, E. P., Adamson, J. J., Ram, K., Poisot, T., & Gravel, D. (2013). The case for open preprints in biology. PLoS Biol, 11(5), e1001563.

Devlin, J., Chang, M. W., Lee, K., & Toutanova, K. (2018). Bert: Pre-training of deep bidirectional transformers for language understanding. arXiv preprint 1810.04805.

Eisen, M. B., Akhmanova, A., Behrens, T. E., & Weigel, D. (2020). Peer Review: Publishing in the time of COVID-19. Elife, 9, e57162.

Epstein, S. (1996). Impure science: AIDS, activism, and the politics of knowledge (Vol. 7). Univ of California Press.

Fan, L., & Yao, Y. G. (2011). MitoTool: a web server for the analysis and retrieval of human mitochondrial DNA sequence variations. Mitochondrion, 11(2), 351–356.

Fraser, N., Brierley, L., Dey, G., Polka, J. K., Pálfy, M., & Coates, J. A. (2020). Preprinting a pandemic: the role of preprints in the COVID-19 pandemic. bioRxiv.

Gale, R. P. (2020). Conquest of COVID-19. Publish it to Death?. British Journal of Haematology.

Gianola, S., Jesus, T. S., Bargeri, S., & Castellini, G. (2020). Publish or perish: Reporting Characteristics of Peer-reviewed publications, pre-prints and registered studies on the COVID-19 pandemic. medRxiv.

Glasziou, P. P., Sanders, S., & Hoffmann, T. (2020). BMJ. Waste in covid-19 research.

Hampton, K. N. (2016). Persistent and pervasive community: New communication technologies and the future of community. American Behavioral Scientist, 60(1), 101–124.

Horbach, S. P. (2020). Pandemic Publishing: Medical journals strongly speed up their publication process for Covid-19. Quantitative Science Studies, 1–12.

Horowitz, M. (2020, May 15). Conflict, Coronavirus, Power, and Security, The Global Cable: Conflict and Competition, and COVID-19 with Michael Horowitz. A Conversation. Perry World House, The University of Pennsylvania. Retrieved from https://global.upenn.edu/perryworldhouse/news/conflict-competition-and-covid-19-michaelhorowitz

Iwema, C. L., LaDue, J., Zack, A., & Chattopadhyay, A. (2016). search. bioPreprint: a discovery tool for cutting edge, preprint biomedical research articles. F1000Research, 5.

Kaiser, J. (2017). Are preprints the future of biology? A survival guide for scientists. Science, 485.

Karr, David. (2020, March 16). Publishers make coronavirus (COVID-19) content freely available and reusable. Wellcome Trust. Retrieved from https://wellcome.ac.uk/press-release/publishersmakecoronavirus-covid-19-content-freely-available-and-reusable

Keck, F. (2020). Asian tigers and the Chinese dragon: Competition and collaboration between sentinels of pandemics from SARS to COVID-19. Centaurus, 62(2), 311–320.

Kirkham, J. J., Penfold, N., Murphy, F., Boutron, I., Ioannidis, J. P., Polka, J. K., & Moher, D. (2020). A systematic examination of preprint platforms for use in the medical and biomedical sciences setting. bioRxiv.

Klein, M., Broadwell, P., Farb, S. E., & Grappone, T. (2019). Comparing published scientific journal articles to their pre-print versions. International Journal on Digital Libraries, 20(4), 335–350.

Kuhn, T. S. (1962). The structure of scientific revolutions. University of Chicago Press.

Kwon, D. (2020). How swamped preprint servers are blocking bad coronavirus research. Nature.

Klebel, T., Reichmann, S., Polka, J., McDowell, G., Penfold, N., Hindle, S., & Ross-Hellauer, T. (2020). Peer review and preprint policies are unclear at most major journals. BioRxiv.

Larivière, V., Sugimoto, C. R., Macaluso, B., Milojević, S., Cronin, B., & Thelwall, M. (2014). arXiv E prints and the journal of record: An analysis of roles and relationships. Journal of the Association for Information Science and Technology, 65(6), 1157–1169.

Leaman, R. (2020). A Comprehensive Dictionary and Variability Analysis of Terms for COVID-19 and SARS-CoV-2.

Lin, J., Yu, Y., Zhou, Y., Zhou, Z., & Shi, X. (2020). How many preprints have actually been printed and why: a case study of computer science preprints on arXiv. Scientometrics, 1–20.

LitCovid. (2020, August 24). Weekly Publications Graph. Retrieved from https://www.ncbi.nlm.nih.gov/research/coronavirus/

Majumder, M. S., & Mandl, K. D. (2020). Early in the epidemic: impact of preprints on global discourse about COVID-19 transmissibility. The Lancet Global Health, 8(5), e627–e630.

medRxiv COVID-19 (2020, August 27th). COVID-19 SARS-CoV-2 preprints from medRxiv and bioRxiv. Retrieved from https://connect.medrxiv.org/relate/content/181

Merton, R. K. (1973). The sociology of science: Theoretical and empirical investigations. University of Chicago press.

McDonnell, D. (2020, April 8). A ‘bridge to a vaccine’: The race to roll out antibody-based Covid-19 drugs. Quartz. Retrieved from https://qz.com/1835197/pharma-companies-race-to-roll-outantibodybased-covid-19-drugs/

medRxiv landing page. (2020). Retrieved from https://www.medrxiv.org/

National Institutes of Health, Office of Portfolio Analysis. (2020). iSearch COVID-19 Portfolio. Retrieved from https://icite.od.nih.gov/covid19/search/

National Institutes of Health Preprint Pilot. (2020). Retrieved from https://www.ncbi.nlm.nih.gov/pmc/about/nihpreprints/

Nature Editorial. (2020, February 4). Calling all coronavirus researchers: keep sharing, stay open. Nature. Retrieved from https://www.nature.com/articles/d41586-020-00307-x

Novaes, M. G., & Guilhem, D. B. (2020). Does the COVID-19 pandemic reveal challenges for open science and the credibility of scientific dissemination?. Revista Brasileira de Farmácia Hospitalar e Serviços de Saúde, 11(2), 0493–0493.

Outbreak Science Rapid PREreview. (20202). Retrieved from https://outbreaksci.prereview.org/

Pastick, K. A., Okafor, E. C., Wang, F., Lofgren, S. M., Skipper, C. P., Nicol, M. R., … & Schwartz, I.. (2020, April). Hydroxychloroquine and chloroquine for treatment of SARS-CoV-2 (COVID-19). In Open Forum Infectious Diseases (Vol. 7, No. 4, p. ofaa130). US: Oxford University Press.

Pirinen, T., & Lindén, K. (2010). Finite-State Spell-Checking with Weighted Language and Error Models:Building and Evaluating Spell-Checkers with Wikipedia as Corpus. In Proceedings of LREC 2010 Workshop on creation and use of basic lexical resources for less-resourced languages.

Rapid Reviews COVID-19 (RR:C19). (2020). Retrieved from https://rapidreviewscovid19.mitpress.mit.edu/

Retraction Watch. (2020). Tracking retractions as a window into the scientific process. Retrieved from https://retractionwatch.com/

Schloss, P. D. (2017). Preprinting microbiology. Mbio, 8(3).

Seatgeek fuzzywuzzy python package. (2020). Retrieved from https://github.com/seatgeek/fuzzywuzzy

Serghiou, S., & Ioannidis, J. P. (2018). Altmetric scores, citations, and publication of studies posted as preprints. Jama, 319(4), 402–404.

Shopovski, J., & Sloboda, B. (2020). Covid-19 Pandemic, the Value of Open Access to Research, and Role of Agile Peer Review. European Scientific Journal, ESJ.

Singh Chawla, D. (2017). When a preprint becomes the final paper. Nature News.

Singh, T., Kumari, M., & Mahajan, S. (2017). Feature oriented fuzzy logic based web spam detection. Journal of Information and Optimization Sciences, 38(6), 999–1015.

Smaglik, P. (1999). E-biomed becomes PubMed Central. The Scientist, 13(19).

Vale, R. D. (2015). Accelerating scientific publication in biology. Proceedings of the National Academy of Sciences, 112(44), 13439–13446.

Varmus, H. (1999). E-BIOMED: A Proposal for Electronic Publications in the Biomedical Sciences, National Institutes of Health. Retrieved from https://profiles.nlm.nih.gov/spotlight/mv/catalog/nlm:nlmuid-101584926X356-doc

Vlasschaert, C., Topf, J., & Hiremath, S. (2020). Proliferation of papers and preprints during the COVID 19 pandemic: Progress or problems with peer review?. Advances in Chronic Kidney Disease.

Yeo-Teh, N. S. L., & Tang, B. L. (2020). An alarming retraction rate for scientific publications on Coronavirus Disease 2019 (COVID-19). Accountability in research, 1–7.

Wang, L. L., Lo, K., Chandrasekhar, Y., Reas, R., Yang, J., Eide, D., … & Mooney, P. (2020). CORD-19: The Covid-19 Open Research Dataset. ArXiv.

Whitley, R. (1984). The social and intellectual organization of the sciences.

